# Comparison of SARS-CoV-2 detection in nasopharyngeal swab and saliva

**DOI:** 10.1101/2020.05.13.20100206

**Authors:** Sumio Iwasaki, Shinichi Fujisawa, Sho Nakakubo, Keisuke Kamada, Yu Yamashita, Tatsuya Fukumoto, Kaori Sato, Satoshi Oguri, Keisuke Taki, Hajime Senjo, Junichi Sugita, Kasumi Hayasaka, Satoshi Konno, Mutsumi Nishida, Takanori Teshima

## Abstract

We prospectively compared the efficacy of PCR detection of SARS-CoV-2 between paired nasopharyngeal and saliva samples in 76 patients including ten COVID-19 patients. The overall concordance rate of the virus detection between the two samples was 97.4% (95%CI, 90.8–99.7). Viral load was equivalent in COVID-19 patients, but the virus tended to disappear earlier in saliva at convalescent phase compared to nasopharyngeal samples. These results suggest that saliva is a reliable noninvasive alternative to nasopharyngeal swabs and facilitate widespread PCR testing in the face of shortages of swabs and protective equipment without posing a risk to healthcare workers.

## Introduction

Rapid detection of the novel coronavirus SARS-CoV-2 is critical for the prevention of outbreaks coronavirus disease 2019 (COVID-19) in communities and hospitals. The diagnosis of COVID-19 is made by PCR testing of samples collected by nasopharyngeal or oropharyngeal swabs, with the nasopharyngeal route being the standard with a sensitivity for the virus in the range of 52–71%(1–5). However, swab sample collection requires specialized medical personnel with protective equipment and poses a risk of viral transmission to healthcare workers. Although sputum specimen is a noninvasive alternative, sputum production is seen in only 28%of COVID-19 patients(6).

The angiotensin converting enzyme 2 (ACE2) is the main host cell receptor for SARS-CoV-2 entry to the human cell(7, 8). ACE2 is highly expressed on the mucous of oral cavity, particularly in epithelial cells of the tongue(9). These findings explain mechanisms that the oral cavity is high risk for SARS-CoV-2 infection, transmission occurs through saliva before the onset of symptoms, and the impairment of the sense of taste(10). Thus, it is reasonable to use saliva as a diagnostic sample, and recent studies have shown that SARS-CoV-2 is detected in saliva(11–13). Moreover, Wyllie et al. demonstrated the saliva to be more sensitive for SARS-CoV-2 detection patients than nasopharyngeal swabs(14). However, few studies compared viral load between nasopharyngeal and saliva samples. We herein compared the diagnostic value of saliva and nasopharyngeal samples using prospectively collected paired samples.

## Methods

### Patients

Nasopharyngeal swab samples and saliva samples were simultaneously collected from patients suspicious of COVID-19 and from patients who were referred to our hospital with the diagnosis of COVID-19. This study was approved by the Institutional Ethics Board and informed consent was obtained from all patients.

### Sample collection

Nasopharyngeal samples were obtained by using FLOQSwabs (COPAN, Murrieta, CA, USA). The swab was passed through the nostril until reaching the posterior nasopharynx and slowly removed while rotating. The swabs were placed in the saline. Saliva samples were self-collected by the patients except one patient, in whom saliva was collected by swab due to inability of self-collection, and spit into a sterile PP Screw cup 50 (ASIAKIZAI Co., Tokyo, Japan). 200 μL Saliva was added to 600 μL PBS, mixed vigorously, then centrifuged at 20,000 X g for 5 minutes at 4°C, and 140 µl of the supernatant was used as a sample.

### PCR

Real-time reverse transcription-quantitative PCR (RT-qPCR) was conducted according to the manual for the Detection of Pathogen 2019-nCoV Ver.2.9.1, March 19, 2020, by the National institute of infectious diseases (https://www.niid.go.jp/niid/images/lab-manual/2019-nCoV20200319.pdf, accessed 2020-5-3). Total RNA was extracted by QIAamp Viral RNA Mini Kit (QIAGEN, Hilden, Germany) from each specimen. One-step RT-qPCR was performed by One-Step Real-Time RT-PCR Master Mixes (Thermo Fisher Scientific, Waltham, USA), from extracted RNA. The instrument used was the StepOnePlus Real Time PCR System (Thermo Fisher Scientific). Forward primer (5-AAATTTTGGGGACCAGGAAC-3), reverse primer (5-TGGCAGCTGTGTAGGTCAAC-3) and TaqMan probe (5’-FAM-ATGTCGCGCATTGGCATGGA-BHQ-3’) were used.

### Statistical analysis

We used the paired t-test to compare data. All *P-values* were 2-sided. Agreement between nasopharyngeal and saliva samples for the detection ability of SARS-CoV-2 was assessed using Cohen’s Kappa(15), presented with 95% confidence interval. All statistical analyses were performed with EZR (Jichi Medical University, Saitama, Japan), which is a graphic user interface for R (The R Foundation for Statistical Computing, Vienna, Austria). *P-value* of 0.05 was used as the cutoff for statistical significance.

## Results

Seventy-six patients were enrolled in this study, including 10 patients with COVID-19 and sixty-six COVID-19 suspicious patients. Most of COVID-19 patients had mild to moderate disease, with no patient requiring ventilator. Median age of COVID-19 patients was 69 years-old, ranging from 30 to 97 years-old. Nine COVID-19 patients were admitted to our hospital after a diagnosis was made by nasopharyngeal samples, while a diagnosis was made at our hospital in one patient. In COVID-19 patients, median day of sampling was 9 days (range, 3–19 days) after symptom onset. SARS-CoV-2 was detected in 8 /10 patients in both nasopharyngeal and saliva samples, and in either nasopharyngeal or saliva sample only in 2 / 10 patients (Table 1). In one patient who showed swab negativity, samples were taken 3 days after symptom onset, while in one patient who showed saliva negativity, samples were taken 19 days after symptom onset. The overall concordance rate of the virus detection was as high as 97.4% (95%CI, 90.8-99.7, Table 1). Concordance between the two samples was strong, as judged by Kohen’ s kappa coefficient(15). The viral loads and the cycle threshold (CT) values defined as the number of cycles required for the fluorescent signal to cross the threshold were not significantly different between nasopharyngeal and saliva samples (Table2). When looking at relation of viral load and duration from symptom onset to sampling, viral load was equivalent between the two samples at earlier time points but declined in saliva at later time points (Figure 1A). The same went for the CT values; they were not significantly different between the two samples but tended to be higher in saliva than in nasopharyngeal samples later (Figure 1B).

**Table 1.**
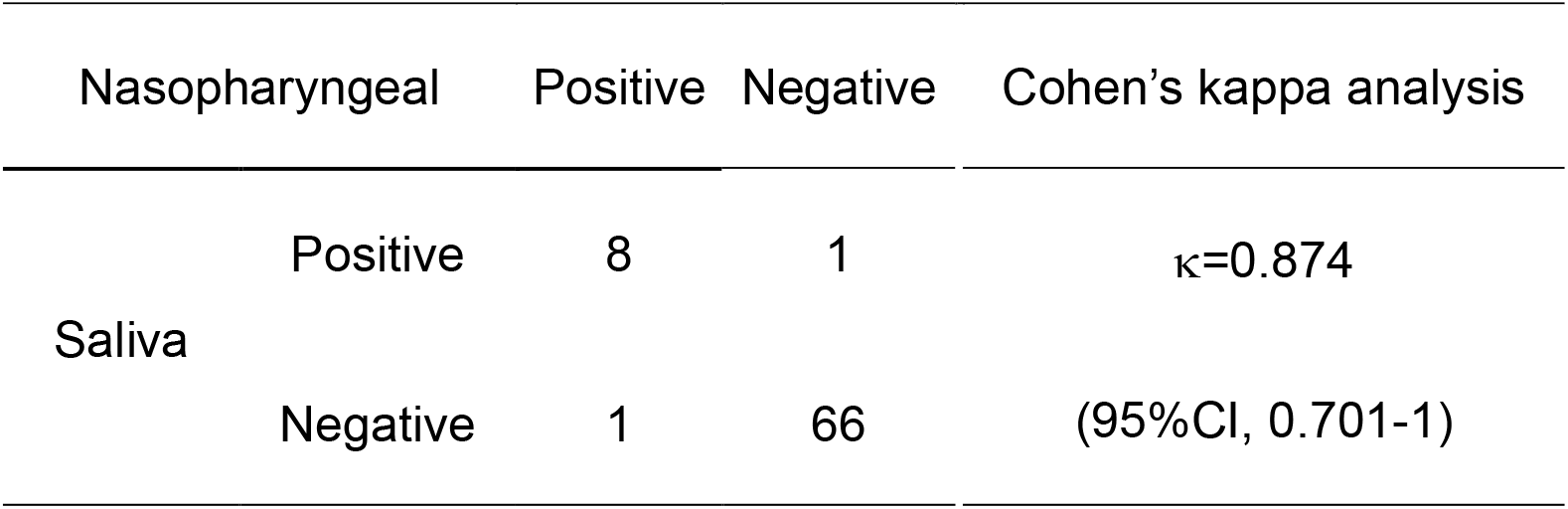
Concordance of PCR results in COVID-19 patients between nasopharyngeal and saliva samples

**Table 2.** Viral roads and cycle threshold values of nasopharyngeal and saliva samples

|  | Nasopharyngeal | Saliva | P value |
| --- | --- | --- | --- |
| Positive | 9/10 (90%) | 9/10 (90%) |  |
| Viral load<br>(Log10 gene copies/mL) | $5.4 \pm 2.4$ | $4.1 \pm 1.4$ | 0.184 |
| Cycle threshold value | $26.5 \pm 8.1$ | $30.6 \pm 4.6$ | 0.206 |

Data are shown as mean ± SD

**Figure 1.**
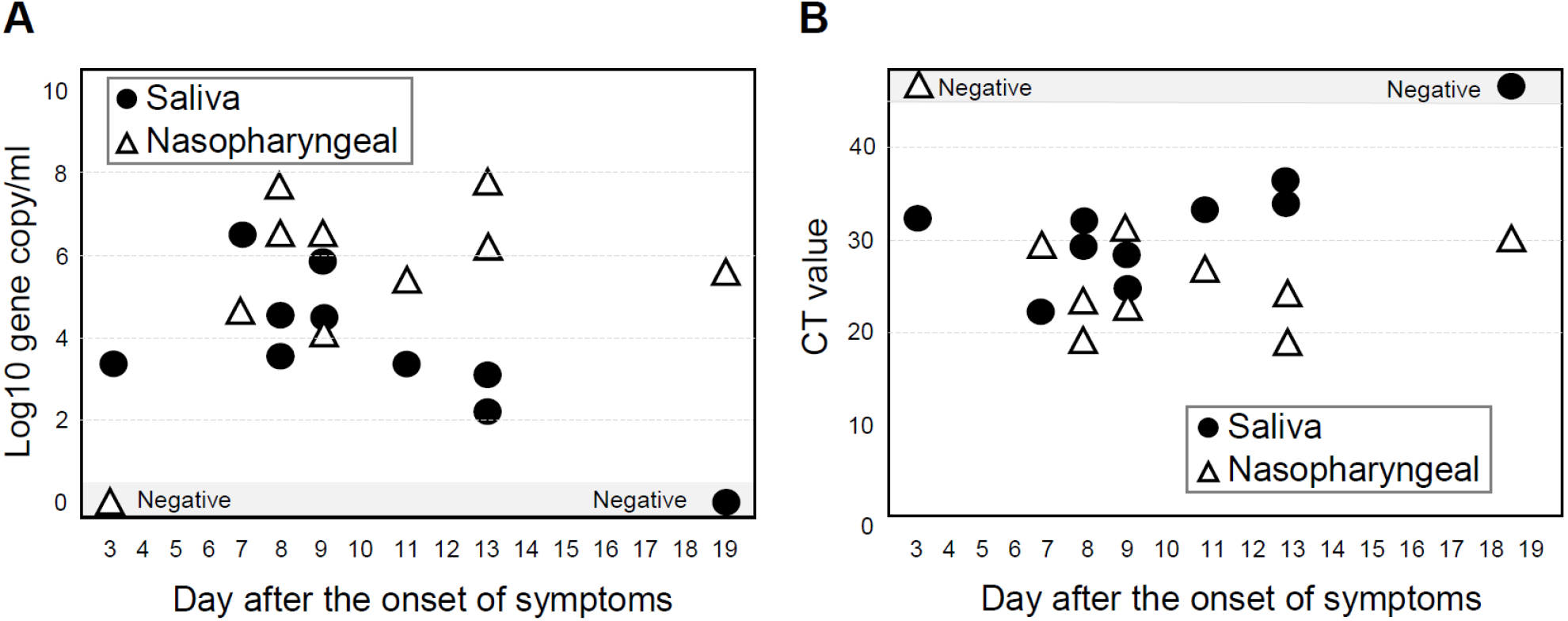
CT values and viral loads according to time after the onset of symptoms. Viral loads (A) and CT values (B) are shown according to time from the onset of symptoms. Closed circle and solid line, saliva sample; Open triangle and dashed line, nasopharyngeal samples.

All patients were treated with favipiravir(16).Figure 2 shows the results of PCR tests in all 28 samples taken from the 10 patients according time from symptom onset in each patient. All 12 saliva samples taken within 2 weeks after COVID-19 onset were positive in saliva. After 2 weeks, PCR negativity was achieved in some of the samples.

**Figure 2.**
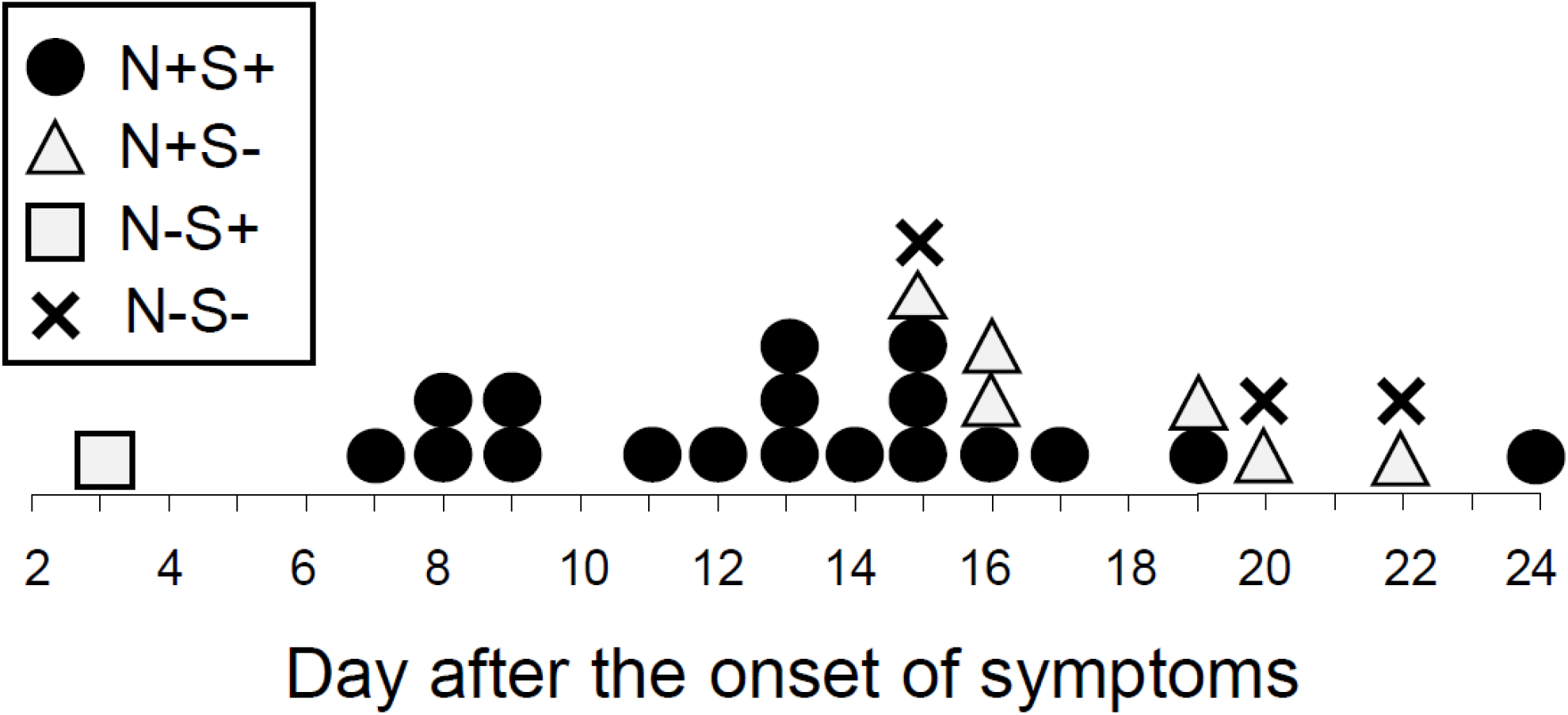
Detection of SAS-CoV-2 in nasopharyngeal and saliva samples. Results of multiple PCR testing from 10 patients shown according to the day after onset of symptoms. N, nasopharyngeal; S, saliva; +, positive;−, negative

## Discussion

We prospectively compared SARS-CoV-2 detection between nasopharyngeal samples and saliva samples in 76 patients. The overall concordance rate of the virus detection was as high as 97.4%. Previous studies also demonstrated high detection rate of the virus in saliva: 11/12 (92%) patients and 20/23 (87%) patients in Hong Kong(11, 13), 25/25 (100%) in Italy(12), and 36/38 (95%) in New Haven(14). In a screening clinic in Australia, 39/622 (6.3%) patients had PCR positive nasopharyngeal swabs and among them, 33/39 (85%) had virus in saliva(17). Taken together, these results consistently support the use of saliva as an effective alternative to nasopharyngeal swabs for diagnosis and screening of COVID-19.

It has been shown that salivary viral load peaks at onset of symptoms and is highest during the first week and subsequently declines with time(11, 13, 14). Our results were consistent to these data; the virus was detected in all the saliva samples taken within 2 weeks after symptom onset. On the other hand, at convalescent phase the viral load decreased earlier in saliva compared to nasopharyngeal samples. These results are consistent to a recent report showing that sensitivity of saliva samples decrease at later time points after symptom onset(18, 19). Recent reports demonstrate that particle of the dead virus could persist in the nasopharynx and resulted in “false positivity”. Interestingly, in our results, the virus tended to become negative much quicker in saliva than in the nasopharynx, suggesting that dead virus particle in mouth is more efficiently cleaned by saliva. Saliva could be a better tool to determine virus clearance in COVID-19 patients.

To our knowledge, four studies compared viral load between nasopharyngeal and saliva samples. The viral loads were 5-times higher in saliva than in nasopharyngeal samples in one study(14), whereas they were lower in saliva in two studies(17, 19). In one study, viral loads were equivalent in symptomatic patients, but lower in asymptomatic patients in saliva(20). Our results showed that the viral load was equivalent at earlier time points but lower in saliva than in nasopharyngeal samples at convalescent phase. Several studies suggested that salivary viral load is highest during the first week after symptom onset and subsequently declines with time(11, 13, 14). Timing of sampling, severity of the disease, different methodologies of saliva collection and processing, different skill of swab sampling may be related to inconsistent results.

Although our study has several limitations due to the small number of samples and few samples within the first week of symptom onset, there have been few prospective studies to date comparing the two samples. Given the large benefits of saliva collection that does not require health worker specialists and protective equipment, our results together with recent studies support the use of saliva as a noninvasive alternative to nasopharyngeal swabs to greatly facilitate widespread PCR testing.

## Data Availability

Data are available.

